# Post-acute sequelae after Nipah virus infection: a systematic review

**DOI:** 10.64898/2026.02.03.26345343

**Authors:** Tiantian Zhang, Qingshuang Wei, Nora Schmit

## Abstract

Incidence patterns of post-acute sequelae, characterised by persistence or delayed onset after the acute phase of an infection, are not well documented after infectious disease outbreaks. Nipah virus was first detected in the 1998-1999 Malaysia outbreak and remains a significant public health concern due to its high epidemic potential and recurrent outbreaks in South Asia. We conducted a systematic review on the prevalence, incidence, duration, and characteristics of post-acute sequelae in survivors of Nipah virus infection. We searched PubMed and Web of Science for studies published up to November 17, 2025. We included 8 articles, and extracted prevalence for 34 potential neurological, psychiatric or non-specific post-acute sequelae. The pooled prevalence of total residual neurological deficits was 24% (95% CI 9-49) among total survivors of Nipah infection, and 45% (95% CI 11-85) among the subset of survivors with acute Nipah encephalitis (5 articles). In the single controlled study, total residual neurological deficits, fatigue and daytime somnolence were significantly more prevalent in Nipah infection survivors than population-based controls. We estimated that 10% (95% CI 4-20) of Nipah infection survivors also experience late-onset or relapsing neurological symptoms after initial recovery. These findings demonstrate a substantial long-term disease burden following Nipah virus infection, which should be accounted for in mathematical modelling studies. However, the estimates are mainly based on data from the Malaysia/Singapore outbreak and may not be generalisable to the Bangladeshi and Indian setting, where current outbreaks occur and are caused by a different viral strain. Further limitations relate to subjective outcome assessment and heterogeneous populations of total Nipah infection survivors, which could have biased our estimates.

## Introduction

The COVID-19 pandemic has highlighted the significant long-term disease burden that can follow infectious disease outbreaks, affecting survivors for months or years after the initial infection.^1^ A fifth of patients were estimated to still experience at least one unresolved symptom 3 years after the acute SARS-CoV-2 infection, such as dyspnea or fatigue.^2^ Such post-acute sequelae or post-acute infection syndromes can also be triggered by other infectious diseases, can affect various organ systems, and include a broad range of persistent or newly emerging symptoms.^1,3^ Waves of post-acute sequelae after several major historical outbreaks have been described in the literature.^1^ However, due to their often heterogeneous and non-specific nature and the lack of objective diagnostic criteria, the extent and similarities in post-acute sequelae following different epidemics have not been systematically documented.

In the World Health Organisation (WHO) 2018 R&D Blueprint, Nipah virus was identified as one of nine priority pathogens with high epidemic potential, high case fatality rate and a lack of approved vaccines and therapeutics.^4^ Nipah virus is a zoonotic virus belonging to the genus *Henipavirus* and was first detected in Peninsular Malaysia in September 1998.^5,6^ The 1998-1999 outbreak resulted from transmission from infected pigs and caused 283 symptomatic cases and 109 deaths, and also spread to slaughterhouse workers in Singapore in 1999.^6^ Since then, recurrent outbreaks have occurred in Bangladesh and India.^7^ The typically small clusters mainly result from consumption of raw date palm sap contaminated by infected *Pteropus* bats, the reservoir of Nipah virus. However, human-to-human transmission has also contributed to these outbreaks, particularly in the healthcare setting.^8^ The bat reservoir has a broad geographical distribution, making new spillover events in other areas likely.^6,9^

The majority of Nipah virus infections are thought to be symptomatic, with the proportion of asymptomatic or mild infections estimated at less than 15%.^10,11^ The main clinical features are acute encephalitis and respiratory disease, with differences between the viral strain originating in Malaysia (Nipah virus-Malaysia strain; NiV-M) and that in South Asia (Nipah virus-Bangladesh strain; NiV-B). While neurological symptoms predominated in the Malaysia/Singapore outbreak, respiratory involvement has more commonly occurred in India and Bangladesh.^6,12,13^ Symptomatic Nipah virus infections usually begin with fever, headache, dizziness and vomiting.^5^ After development of encephalitis, patients can deteriorate to coma and death within days,^6^ with a high case fatality rate of around 61% which varies by country.^14^ Among survivors, instances of neurological relapse and residual neurological deficits after the acute infection have been reported since the first patient descriptions.^15^

Synthesising information on pathogen epidemiology is critical for epidemic preparedness, and a better definition of disease burden has been identified as a strategic goal among key research priorities for the development of Nipah virus medical countermeasures.^9^ Estimates of key transmission and disease parameters can inform parameterisation of mathematical models for future outbreak response and intervention evaluation; a collection of systematic reviews on the epidemiology of the 2018 WHO priority pathogens is ongoing.^16-19^ Here, we complement this by synthesising the currently fragmented evidence base on the potential long-term health effects of Nipah virus infection. We aimed to systematically review and quantify the prevalence, incidence, duration and other characteristics of post-acute sequelae in Nipah virus infection survivors.

## Methods

### Search strategy and selection criteria

This systematic review was conducted according to the PRISMA guidelines^20^ and registered with PROSPERO (registration number: CRD42024616198). A systematic search was conducted in the PubMed and Web of Science databases to identify all available articles on post-acute sequelae associated with Nipah virus, published from database inception to 17 November 2025. The keywords were: (“Nipah virus” OR henipa*) AND (chroni* OR long-term* OR long-standing* OR enduring* OR post* OR sequel* OR surviv* OR recov* OR convalescen* OR disability OR morbidity OR persist* OR rehabilitat* OR complicat* OR aftermath OR residual* OR late-onset OR delayed OR relaps*). The retrieved citations were imported into Covidence software and duplicates were removed.^21^ Articles were screened independently by two reviewers, and discrepant classifications were resolved through discussion with the third author.

Articles were eligible for inclusion if they reported primary data on post-acute sequelae in survivors of Nipah virus infection, without restrictions on study design. Our definition of post-acute sequelae included (a) symptoms persisting for at least 3 months after the initial infection, (b) new symptoms developing after the acute phase of the infection (late-onset symptoms), or (c) recurring symptoms after the acute phase of the infection following a period of remission (relapsing symptoms). To meet inclusion criteria, an article therefore needed a minimum follow-up period of 3 months after acute infection, report late-onset or relapsing symptoms occurring after recovery, or both. We included study populations with clinical Nipah virus infection as well as based on seropositivity alone. Exclusion criteria were articles in languages other than English, non-peer-reviewed publication types (e.g. conference proceedings), non-primary research (e.g. reviews), studies on functional outcomes alone (e.g. disability scales), and case reports.

### Data extraction and analysis

A data extraction template was designed and piloted. Two reviewers independently extracted data on study design, study population, recruitment, methods for diagnosis of infection and sequelae, sample size, participant characteristics, the time at measurement of post-acute sequelae since the acute infection, prevalence and incidence of all post-acute sequelae among Nipah virus survivors and of the same symptoms among controls (where applicable), characteristics of post-acute sequelae (e.g. duration), and risk factors. Risk of bias assessment was conducted using Joanna Briggs Institute (JBI) critical appraisal tools.^22,23^ To reflect the variety of study designs in this review, the tools for prevalence studies, cohort and case-control studies were combined and adapted to include all relevant quality criteria (Table S1). The quality assessment was conducted by two reviewers, and disagreements were resolved through discussion.

All analyses were conducted in *R* version 4.2.2.^24^ Analysed outcomes included the prevalence of overall post-acute neurological deficits in Nipah survivors, the prevalence of other post-acute sequelae, the association between Nipah virus infection and development of post-acute sequelae, risk factors for post-acute sequelae, and the incidence of late-onset or relapsing symptoms. Individual post-acute sequelae were renamed for analysis if they formed part of a group of symptoms reported in multiple studies (e.g. dysmetria is a type of ataxia). They were also broadly categorised into neurological symptoms (cognitive dysfunction, motor or sensory dysfunction, and other), psychiatric symptoms, or systemic and other symptoms.

Random-effects meta-analysis was considered for all outcomes reported in at least two studies (Supplementary Materials p.4). We aimed to estimate outcomes in all survivors of Nipah virus infection, but several included studies only focused on patients hospitalised with acute Nipah encephalitis. We therefore conducted subgroup analyses for these different study populations. Where available, pooled estimates were calculated for (a) total survivors of Nipah infection (regardless of the presentation during acute infection), (b) survivors of Nipah encephalitis, who experienced encephalitis symptoms during acute infection and represent a subset of total survivors of Nipah infection, and (c) survivors of late-onset or relapsed Nipah encephalitis. The latter includes patients who survived both the initial Nipah virus infection (with or without encephalitis) and a subsequent episode of late-onset or relapsed encephalitis. Multiple articles included in the systematic review followed the same or overlapping patient populations (Figure S1). To avoid inclusion of overlapping data in pooled prevalence estimates among Nipah encephalitis survivors, we only included one of the articles in the primary meta-analysis and investigated the effect of including alternative articles in sensitivity analyses. The decision on article inclusion was based on larger sample size in the Malaysian studies, and on alignment with other studies on the definition of residual neurological deficits in the Singaporean studies.

Prevalence estimates for individual post-acute sequelae only reported in one study were summarised descriptively, with 95% confidence intervals calculated using the Clopper-Pearson method. The association between post-acute sequelae and Nipah virus infection was assessed using Fisher’s exact test due to the small sample size in the included study. For all other outcomes, descriptive statistics and narrative synthesis are presented.

## Results

Our search identified 1,091 articles after deduplication, of which 1,017 were excluded in title-and-abstract screening (Figure 1). For 66 articles excluded in full-text screening, the most common exclusion reason was not reporting on post-acute sequelae. Two of the excluded studies described residual neurological deficits, but had a too short follow-up period to meet our case definition (Supplementary Materials p.5).^25,26^

**Figure 1.**
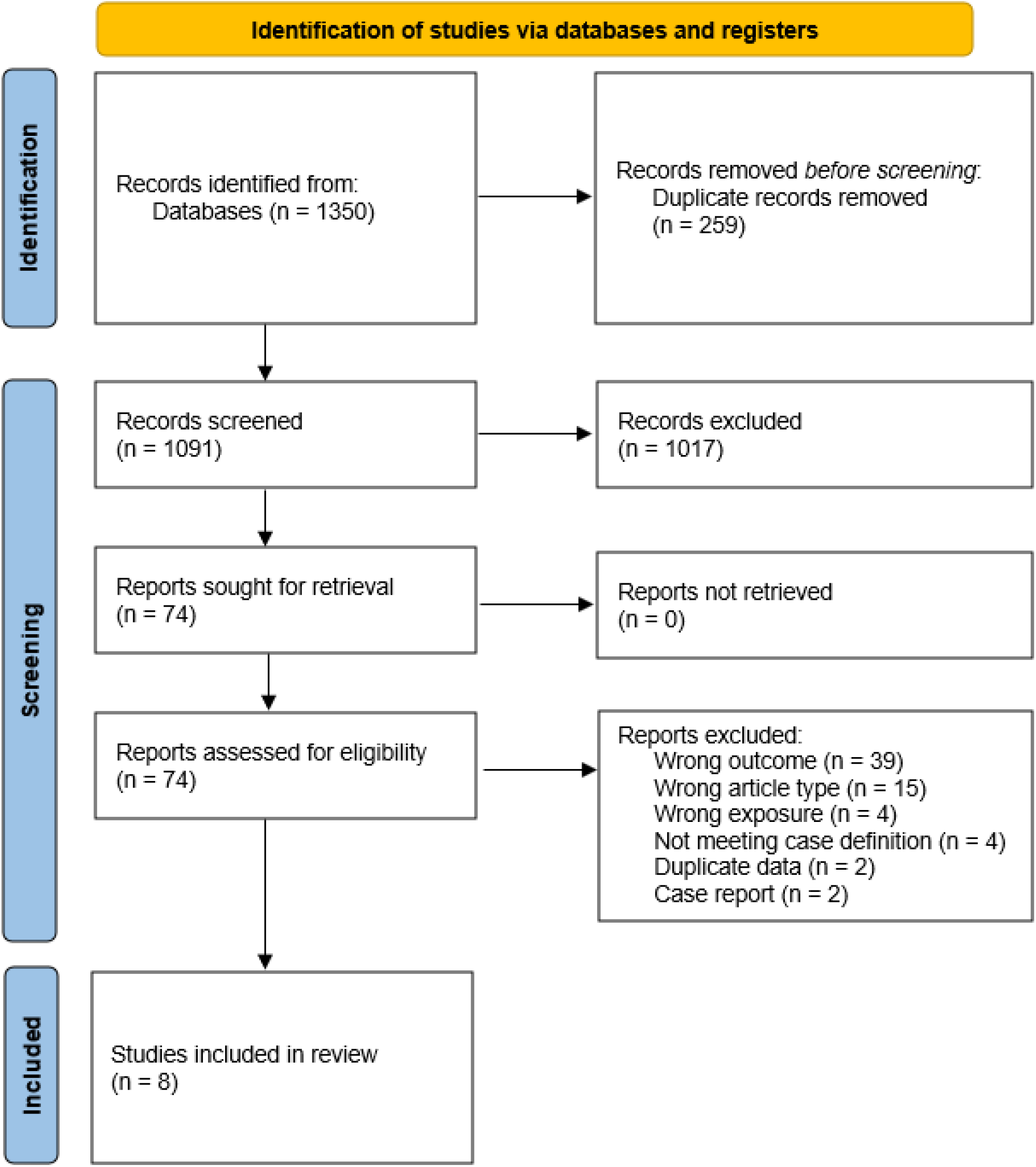
PRISMA flowchart.

### Description of studies

We included 8 articles in the final review.^15,27-33^ Of note, seven articles followed Nipah survivors of the 1998-1999 outbreak in Malaysia and Singapore, while only one was set in Bangladesh and none in India (Table 1). Most studies employed prospective longitudinal designs, with follow-up periods ranging from 3 months to 25 years after acute infection. The main outcomes were neurological sequelae, and some studies also investigated general clinical features; assessment methods included physical/neurological examination, symptom questionnaires, psychiatric evaluation, and neuropsychological testing. In 3 articles (38%) the study population consisted of hospitalised Nipah encephalitis survivors, while the other 5 articles (62%) included survivors of Nipah virus infection more broadly (with encephalitic, non-encephalitic, and asymptomatic acute infection). Only one study included a healthy control group, consisting of household members of Nipah infection survivors who were asymptomatic during the outbreak and seronegative for IgM and IgG antibodies. Sample sizes of Nipah survivors in each article ranged from 9 to 249, though some of these represent the same participants (Supplementary Materials p.3).

**Table 1.** Characteristics of included studies. (*) Applies to Nipah survivors unless otherwise specified.

| First author (year) | Location | Study/ recruitment period | Study design | Patient population | Time at measurement after acute infection/ follow-up | Sample size* | Age* | Sex* | Control group |
| --- | --- | --- | --- | --- | --- | --- | --- | --- | --- |
| **1998-1999 outbreak in Malaysia and Singapore** |  |  |  |  |  |  |  |  |  |
| Goh (2000) | Malaysia | 1999 | Prospective uncontrolled cohort study | Nipah virus encephalitis survivors hospitalised at the University of Malaya Medical Centre (consecutive recruitment) | Unclear (known follow-up until 11 weeks) | 64 | Mean 35 years | 82% male in total patient cohort | / |
| Chong (2002) | Malaysia | 1998-1999 | Retrospective uncontrolled cohort study | All Nipah encephalitis survivors treated in Seremban Hospital | 5 months | 61 | Mean 38 years (SD 14, range 4-75) in total patient cohort | 88% male in total patient cohort | / |
| Tan (2002) | Malaysia | 1998-1999 | Prospective uncontrolled cohort study | Nipah virus infection survivors followed up at various treatment centres in Malaysia | 2 years | 249 | Mean 29 years (range 9-55) in patient cohort with late-onset or relapsed encephalitis | 64% male in patient cohort with late-onset or relapsed encephalitis | / |
| Lim (2003) | Singapore | 1998-1999 | Prospective uncontrolled cohort study | All Nipah encephalitis survivors of the Singapore outbreak, and serologically confirmed abattoir workers with asymptomatic Nipah virus infection | 1.5 years | 21 | Mean 43 years (range 24-65) in encephalitis survivors, mean 50 years in asymptomatic survivors | 95% male | / |
| Ng (2004) | Singapore | 1998-1999 | Prospective uncontrolled cohort study | All Nipah encephalitis survivors of the Singapore outbreak | 2 years | 9 | Mean 41 years (SD 11, range 24-55) | 89% male | / |
| Siva (2009) | Malaysia | 2008-2009 | Retrospective cohort/cross-sectional study | Serologically confirmed Nipah virus infection survivors identified during the outbreak by the University of Malaya, Kuala Lumpur | 10 years | 39 | Mean 46 years (SD 2) | 79% male | 31 seronegative household members of Nipah infection survivors |
| Ong (2025) | Malaysia | 2023 | Retrospective cohort/cross-sectional study | Survivors with a clinical history of Nipah virus infection during the 1998 outbreak from Kampung Sungai Nipah | 25 years | 25 | Mean 39 years (SD 12, range 14-64) | 80% male | / |
| **Other outbreaks** |  |  |  |  |  |  |  |  |  |
| Sejvar (2007) | Bangladesh | 2005-2006 | Retrospective uncontrolled cohort/cross-sectional study | All serologically confirmed symptomatic Nipah virus infection survivors identified during outbreaks in 2003, 2004 and 2005 in Bangladesh | May 2006 (around 23 months after acute infection on average) | 22 | Median 15 years (range 6-50) | 55% male | / |

Three studies (38%) met at least 75% of risk of bias assessment criteria (high quality) and an additional 4 (50%) met at least 50% of criteria (moderate quality) (Table S2). Risk of bias mainly resulted from lack of valid, standard and/or reliable measurement methods for post-acute sequelae (n=5, 63%), lack of confirmation of absence of post-acute symptoms before infection (n=5, 63%), inappropriate sampling (n=4, 50%) and unspecified response rates (n=4, 50%). In Goh et al, outcomes and the follow-up duration were not clearly reported, resulting in ambiguities in the data extraction. Prevalence estimates from this study were excluded from the primary analysis as alignment with our case definition for post-acute sequelae (minimum 3 months persistence) could not be confirmed.

### Prevalence of post-acute sequelae after Nipah infection

#### Residual neurological deficits

Four independent studies published across 6 articles reported the prevalence of total residual neurological deficits. The pooled prevalence among total survivors of Nipah infection was estimated at 24% (95% CI 9-49). In comparison, the prevalence among the subset of Nipah encephalitis survivors was higher (*p* = 0.007), at 45% (95% CI 11-85) (Figure 2). The prevalence of residual neurological deficits in a single study on survivors of relapsed or late-onset encephalitis was similar to the latter at 61% (95% CI 36-83). Data among encephalitis survivors displayed heterogeneity, while the prevalence among total Nipah infection survivors was relatively consistent between studies. Nevertheless, the underlying study populations of total Nipah infection survivors were highly heterogeneous, with the percentage of the population with acute encephalitis ranging from 31-81%. The funnel plot was largely symmetrical but suggests the possibility of small studies with low prevalence not being published (Figure S2). However, the number of included studies was too small to accurately assess publication bias.

**Figure 2.**
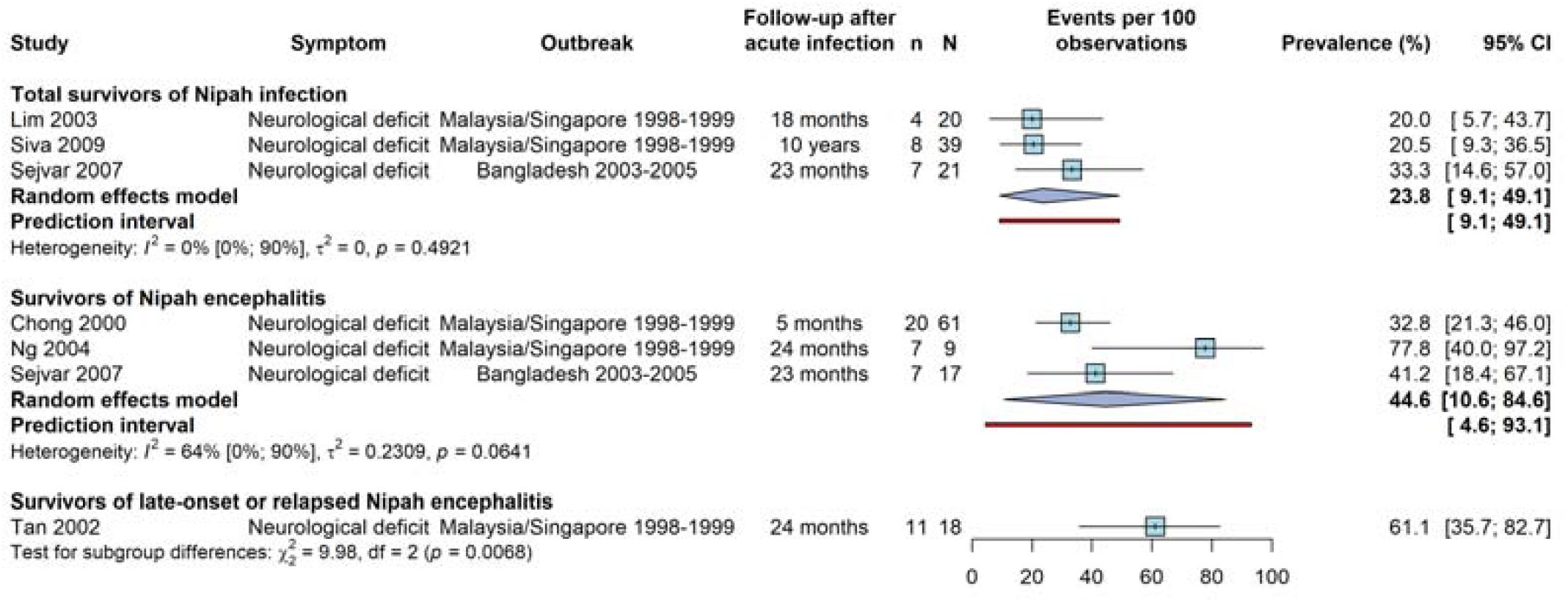
Random-effects meta-analysis for the prevalence of residual neurological deficits after infection with Nipah virus. Prevalence estimates are shown for total survivors of Nipah infection, survivors of Nipah encephalitis, and survivors of late-onset or relapsed Nipah encephalitis (one study). The latter two groups represent a subset of all survivors of Nipah infection. Among Nipah encephalitis survivors, Ng 2004 was included over Lim 2003 due to the inclusion of cognitive impairments among neurological deficits (consistent with the other studies). Chong 2000 was included over Siva 2009 due to a larger sample size. Extraction decisions are detailed in Table S3.

In sensitivity analyses varying the included studies with overlap in study populations (for Nipah encephalitis survivors), results were similar overall (Figure S3). The pooled point prevalence of neurological deficit in Nipah encephalitis survivors ranged from 35% to 59%, but confidence intervals were wide and overlapped in all analyses. When including the study with an unclear follow-up period, the pooled prevalence was 38% (95% CI 15-68) (Figure S4).

All cases of residual neurological deficits among total Nipah infection survivors occurred among encephalitis patients. Based on three studies,^29,31,33^ survivors who experienced acute Nipah encephalitis had a numerically higher prevalence of persistent neurological deficit than those with non-encephalitic acute infection [pooled prevalence ratio 10.0 (95% CI 0.5-183.7)] (Figure S5). Study-specific prevalence ratios ranged from 3.9 to 37.4, but only the study with the largest sample size found a significantly higher prevalence among encephalitis compared to non-encephalitic infection survivors. Sejvar et al. further identified empiric therapy during acute infection as a statistically significant risk factor for residual neurological deficits among survivors.^33^

#### Other symptoms

One study estimated the combined prevalence of any post-acute sequelae persisting more than 1 year after the acute infection at 24% (95% CI 9-45) of 25 Nipah infection survivors (including lethargy, impaired vision, paralysis and seizures).^32^ Of 34 individually reported symptoms in the included studies, the majority (n=24, 71%) were neurological, with 18 classified as motor or sensory dysfunction, 2 as cognitive dysfunction and 4 as other neurological (Figure 3). Motor and sensory dysfunction symptoms affected the limbs, face, eyes, hands, or whole body. The remaining symptoms were categorised as either psychiatric (n=3, 9%) or systemic/other (n=7, 20%). Prevalence for 8 sequelae (24%) was reported in more than one study, namely cognitive impairment, ataxia, cranial nerve palsy, nystagmus, monoparesis, dysarthria, focal weakness, and fatigue (Table S4). The symptoms affecting most patients were fatigue for total Nipah infection survivors [48% (95% CI 23-74) across two studies] and cognitive impairment for Nipah encephalitis survivors [46% (95% CI 14-82) across 2 studies). However, heterogeneity in these studies was high (Table S4). Other frequent neurological symptoms included memory/concentration impairment, nystagmus, bradykinesia, seizures/epilepsy and myoclonus (Figure 3, Table S5). Three studies additionally reported the symptom distribution by patient. Of 27 patients with post-acute sequelae, 67% experienced at least 2 symptoms, with the maximum number of symptoms reported for one patient being 5, and 41% experienced symptoms in multiple categories.

**Figure 3.**
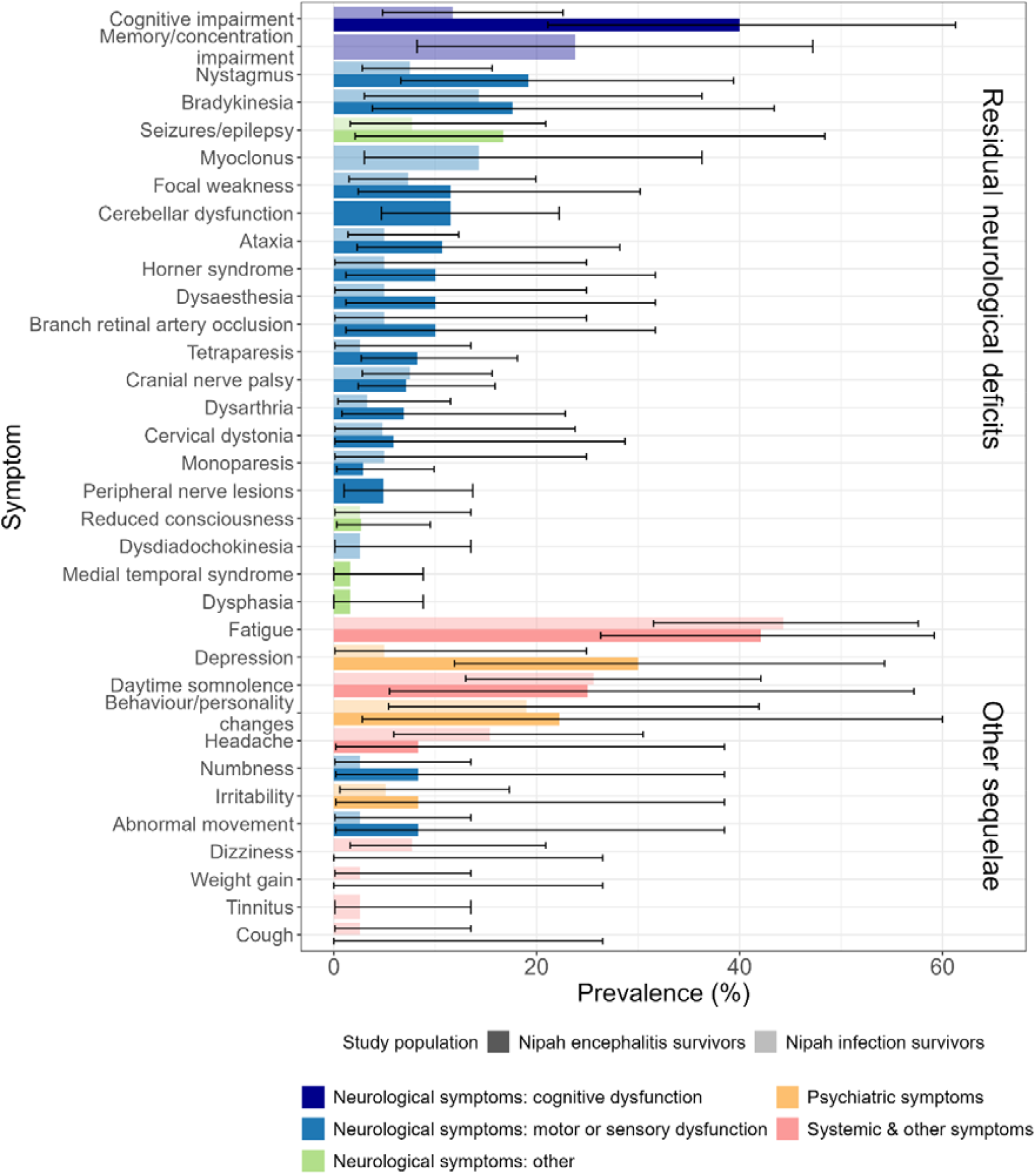
Prevalence for all individual post-acute sequelae reported among total Nipah infection survivors and Nipah encephalitis survivors. The top 22 symptoms form part of the aggregated residual neurological deficits estimate. For symptoms reported in at least two independent studies, prevalence represents the weighted percentage from random-effects meta-analysis (for cognitive impairment, ataxia, cranial nerve palsy, nystagmus, monoparesis, dysarthria, focal weakness, and fatigue). For estimates from different non-independent articles, the mean is shown. Fatigue includes symptoms described as “fatigue/lethargy”, “disabling fatigue lasting more than 6 months” and “chronic fatigue syndrome”. Focal weakness includes facial and finger weakness.

### Association of post-acute sequelae with Nipah

Only one article compared the prevalence of post-acute sequelae among survivors of Nipah infection with a healthy control group, at 10 years after the acute infection.^31^ Residual neurological deficits, fatigue and daytime somnolence were more common in 39 Nipah survivors than the 31 household controls, but there was no evidence for a statistically significant difference for 11 other symptoms (Figure S6).

### Symptom prevalence over time and duration of post-acute sequelae

Persistent neurological and psychiatric symptoms were reported from within 6 months after acute infection^27^ to as late as 10 years after the acute infection.^33^ There were differences in prevalence between studies estimating this at different time points after acute infection (Figure 4). Among total survivors of Nipah infection, the median prevalence for all individual neurological and psychiatric symptoms was lower when measured at 10 years [3% (range 3-8%)] compared to earlier timepoints [5% (range 5-23%) at 1-1.5 and 14% (5-24%) at 1.5-2 years]. Among survivors of Nipah encephalitis, the median prevalence was also highest at 1.5-2 years after acute infection [11% (range 5-75%)], and similar at 0.25-0.5, 1-1.5 and 10 years after acute infection [3.5% (range 2-11%), 5% (range 5-18%) and 3% (3-8%), respectively]. Systemic/other symptoms were only reported at 0.25-0.5 and 10 years after acute infection.

**Figure 4.**
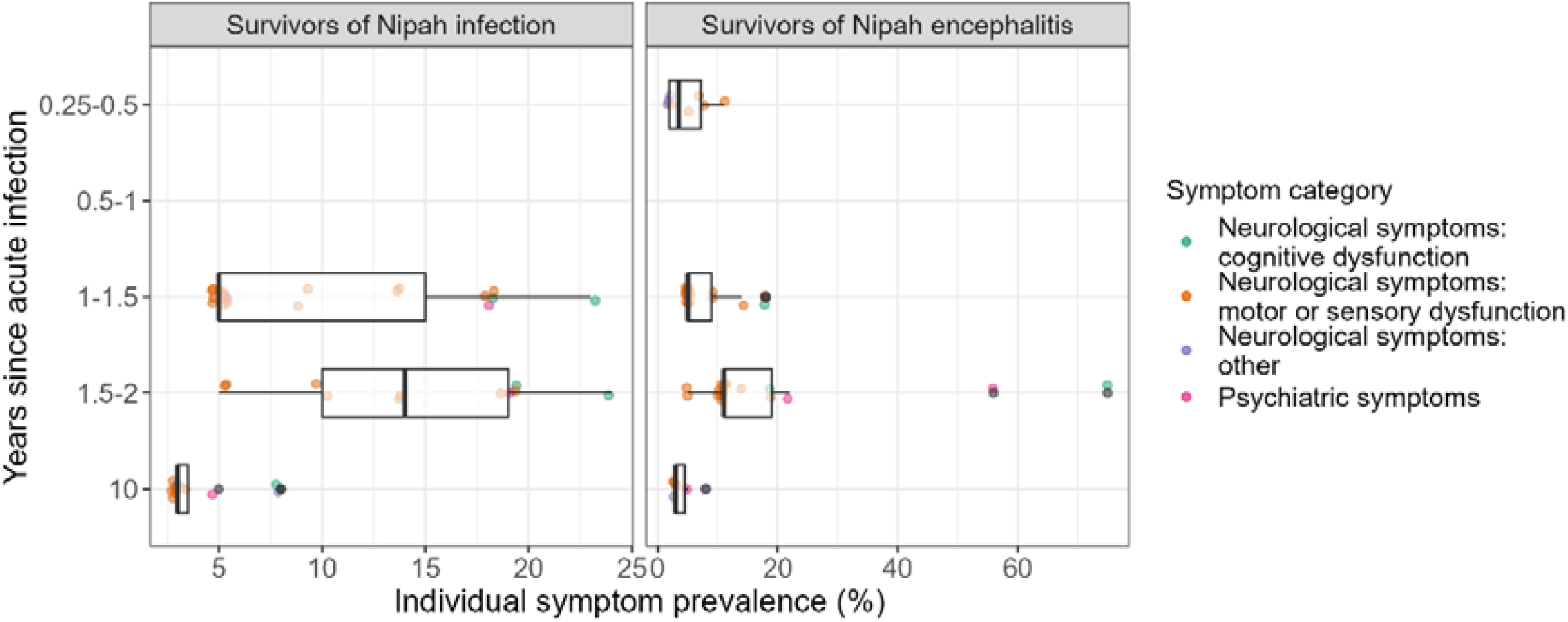
Boxplot of persistent neurological and psychiatric symptom prevalence over time. Points show the individual symptom prevalence datapoints. The plot includes estimates from overlapping study populations if they were measured in different time categories.

In Ong et al., 33% of 9 Nipah infection survivors with post-acute sequelae recovered within 1 year of the acute infection.^32^ In other studies, there was no change in individual symptom prevalence between 14 and 23 months^33^ and between 18 and 24 months.^29,30^ The prevalence of fatigue among survivors of Nipah infection in Sejvar et al. was strongly dependent on time at measurement, declining from 68% at 6 months to 14% at 2 years.^33^ For those recovering within 2 years, the median duration of fatigue was 5 months (range 8 days to 8 months).

### Incidence of late-onset or relapsing neurological sequelae

Three articles estimated the cumulative incidence of late-onset or relapsing neurological symptoms (neurological deficits or encephalitis) after the initial Nipah infection over follow-up periods of 1.5-2 years. In a meta-analysis, there was no significant difference in cumulative risk between overall infection and encephalitis survivors, with estimates of 10% (95% CI 4-20) and 12% (95% CI 2-49), respectively (Figure 5). There was evidence for between-study heterogeneity among Nipah encephalitis survivors (*I*^*2*^ = 60%), with a potentially lower risk of late-onset or recurring encephalitis than neurological deficits. In Tan et al., 54% of late-onset or relapsing Nipah encephalitis patients had encephalitis during the acute infection, 23% had symptomatic non-encephalitic acute infection, and 23% had asymptomatic acute infection.^28^ In Sejvar et al., 100% of delayed-onset neurological abnormalities were among encephalitis survivors.^33^ Goh et al. also reported 4 cases of delayed-onset sequelae or relapse in initially asymptomatic or non-encephalitic survivors, though the denominator was not known.^15^

**Figure 5.**
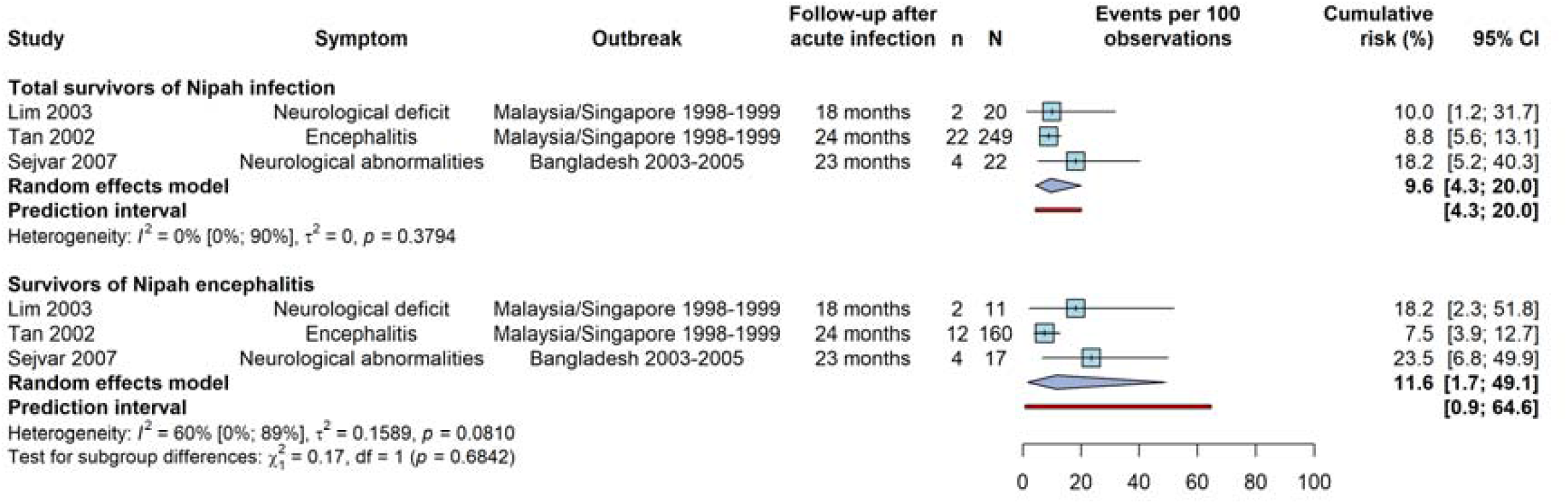
Random-effects meta-analysis for the cumulative risk of late-onset or relapsing neurological sequelae after infection with Nipah virus. Estimates are shown for two subgroups: total survivors of Nipah infection and survivors of Nipah encephalitis (a subset of all survivors of Nipah infection).

All cases of delayed neurological symptom onset with known timing occurred within one year of acute infection (Figure S7). For late-onset or relapsing encephalitis, 50% of 22 patients experienced the first episode by 8.4 months after acute infection, and 14% experienced an additional second episode on average 7.6 months after the first.^28^ For other symptoms, 2 of 5 cases of major depressive disorder had a delayed onset around 1 year after the acute infection.^30^

## Discussion

In this systematic review, we identified 8 studies on post-acute sequelae after Nipah virus infection. A total of 34 individual persistent symptoms were reported among Nipah survivors, mostly systemic (e.g. fatigue) or affecting the nervous system. The results indicate that most Nipah survivors recover without long-term neurological dysfunction. Nevertheless, the pooled prevalence of total residual neurological deficits was substantial and higher among patients with encephalitis during the acute infection, at 45% (95% CI 11-85), than among Nipah survivors overall, at 24% (95% CI 9-49). Total residual neurological deficits, fatigue and daytime somnolence were the only sequelae found to have significantly higher prevalence in Nipah survivors compared to household controls, but we only identified one study with a control group. Further, the results highlight the possibility of late-onset and relapsing symptoms after Nipah infection, with around 10% of survivors estimated to develop new neurological symptoms after recovery from acute infection. Of note, 7 of the 8 included articles focused on the Malaysia/Singapore outbreak, highlighting a lack of published research on sequelae for the Nipah virus-Bangladesh strain that causes current and, likely, future outbreaks. Our estimates may therefore not be generalisable to this strain and the South Asian setting, with known differences in acute clinical presentation and case fatality ratio compared to cases in Malaysia and Singapore.Central nervous system infection is the hallmark complication of acute Nipah virus infection and associated with high mortality.^6^ Neurological sequelae may result from permanent neuronal damage induced by viral infection of the brain or from immune-mediated injury.^29,30^ Cognitive impairment and motor and sensory dysfunction mimic the sequelae seen after other causes of viral encephalitis,^34^ with particularly severe neurological deficits known to occur in Japanese encephalitis survivors.^35^ The observed neurological deficits after Nipah virus infection include many different outcomes of varying severity, from potentially milder symptoms like numbness to being in a vegetative state. The associated functional impairment was generally described as mild, though most patients had some degree of disability (see summary in Table S6). The proportion of Nipah encephalitis survivors with neurological deficits requiring caregiver assistance within 5 months of the acute infection was 15% and 50%,^15,27^ and remained at 15% 10 years after the acute infection.^31^ This suggests that Nipah virus outbreaks may not just incur a high case-fatality rate, but also long-term disability in a subset of patients. Incorporating this in mathematical modelling studies is essential for public health risk assessment and health economic analyses of potential interventions, though estimating the associated health loss is challenging. The disability weight for mild motor plus cognitive impairments (0.054) in the Global Burden of Disease study may reflect some of the less severe outcomes observed in survivors.^36^

A further notable sequela reported in three studies was the high prevalence of fatigue (up to 48% in meta-analyses) both in Malaysian and Bangladeshi survivors. Fatigue was generally characterised as severe and disabling, and found to be significantly more frequent among Nipah infection survivors than population-based controls 10 years after the acute infection.^31^ Severe fatigue is recognised as a common post-infectious sequela after viral and other infections such as SARS-CoV-2 and dengue, which may originate from immune dysregulation triggered by the acute infection.^1,3^ Sejvar et al. noted that all patients with fatigue were unable to resume daily activities for months, highlighting its significant impact on livelihoods and quality of life. However, fatigue appears to resolve within 8 months of infection in the majority of affected patients, persisting long-term only in a smaller subset of survivors.^33^ In contrast, significant neurological deficits were still detected in a similar proportion of Nipah infection survivors at 10 years after acute infection as at earlier timepoints.

Acute disease severity is a known risk factor for the development of post-acute sequelae after SARS-CoV-2 infection.^37^ While direct evidence on an association was limited by small sample sizes, the combined findings suggest that survivors of acute Nipah encephalitis may also be at higher risk of persistent neurological deficits than those with milder acute infection. In the included studies, no significant persistent neurological deficits were detected among a total of 41 asymptomatic or non-encephalitic febrile survivors. Nevertheless, similar brain lesions seen in asymptomatic and encephalitis survivors could indicate the potential for subclinical brain infection by the Nipah virus even in the absence of encephalitis.^29^ Findings from one study also point to the possibility that empiric treatment during the acute infection, rather than the infection itself, may contribute to the development of persistent neurological deficits.^33^ However, this included highly variable therapies (the antiviral acyclovir, antibiotics and corticosteroids). Based on two articles, more than half of Malaysian patients received the antiviral ribavirin during hospitalisation, but the relationship to persistent neurological deficits was not investigated.^15,27^ The Singaporean studies did not describe treatment during the acute infection. We identified no other information on risk factors for developing post-acute sequelae among survivors.

Our findings confirm the potential for late-onset or relapsing neurological sequelae as a characteristic feature of Nipah virus infection. Most of these episodes appear to develop within a year of acute infection (though follow-up in the included studies was limited to 2 years), and they may also occur after mild or asymptomatic acute infections. A few case reports of potential late-onset encephalitis with much longer delays (up to 11 years after the outbreak) have also been published.^38,39^ Similar delayed-onset neurological illness has been observed years after the initial infection with other paramyxoviruses like Hendra virus and measles.^40,41^ Subacute sclerosing panencephalitis is a rare complication due to persistent brain infection with the measles virus,^41^ but there is no direct evidence for Nipah virus persistence in the human central nervous system to date.^28,42^

Important knowledge gaps remain about post-acute sequelae after Nipah virus infection. Firstly, all but one of the included studies related to the Malaysian strain of Nipah virus. We could not identify any published data on patients in India or recent cases in Bangladesh (the included study covered survivors infected in 2003-2005). The prevalence of residual neurological deficits in the study set in Bangladesh was not noticeably different from other estimates,^33^ but the clinical presentation during acute infection and the case fatality ratio is known to differ by country. The available evidence was insufficient to gain strain-specific insights, and so it is not possible to conclude whether post-acute sequelae in Bangladesh and India include respiratory manifestations or otherwise differ from those seen after the Malaysia outbreak. Further, data were not sufficient to estimate the average duration of sequelae, and recovery trajectories remain incompletely characterised. There were also significant methodological limitations in the included studies, as all but one were uncontrolled. The controlled study used household controls, which may not provide a representative estimate of sequelae prevalence in the general population. Outcome assessment methods were also heterogeneous across studies and often subjective, resulting in significant potential for bias in our estimates. However, improving the accuracy and precision of estimates through further research is challenging due to the small numbers of known Nipah virus cases: as of May 2024, there were an estimated 754 reported symptomatic cases, of which 319 survivors, globally.^43^ Almost all of these survivors have been identified in Malaysia (174) and Bangladesh (99). In comparison, our prevalence estimate for encephalitis survivors included 87 individuals (of which 61 from Malaysia).

Our study had several additional limitations. Firstly, our pooled prevalence estimates of residual neurological deficits were highly uncertain and should be interpreted with caution because definitions varied across studies. Siva et al. and Sejvar et al. specifically focused on “significant” and “moderate-to-severe” cases, while Lim et al. did not assess for cognitive impairment in their estimate. Some extracted numbers may also be inaccurate or misinterpreted due to unclear reporting (see Table S3). Additionally, our estimate in total Nipah infection survivors is likely less reliable than that in Nipah encephalitis survivors, as it is not clear how representative the included participants were. This depends, among other factors, on the setting-specific extent of surveillance for mild and asymptomatic infections, treatment standards and healthcare access. While Lim et al. included almost all Nipah survivors identified in mass serological screening in the Singapore outbreak, sampling in the other two studies was not clearly described. The percentage of total Nipah infection survivors with acute encephalitis varied widely from 31% to 81% in the three studies, indicating highly heterogeneous populations.^29,31,33^ Secondly, exclusion of grey literature and non-English-language articles could have introduced publication and language bias in our review. Thirdly, the type of identified post-acute sequelae may be affected by reporting bias, as most studies were conducted by neurologists and focused on neurological outcomes. Lastly, our study-specific definition of post-acute sequelae, with a minimum 3-month persistence, means that some data may have been excluded because follow-up time was not clearly reported or slightly too short.

Despite these limitations, this systematic review provides the first comprehensive assessment of post-acute sequelae after Nipah virus infection. We found that a substantial proportion of survivors experience long-term neurological deficits and fatigue, particularly after Nipah encephalitis. The existing evidence base further suggests that patients recovering from Nipah virus infection should be monitored for the development of late-onset or relapsing neurological symptoms for at least a year, irrespective of their initial presentation. Our estimates and their associated uncertainty should be considered for inclusion in modelling projections of public health risk. However, they need to be interpreted within the context of methodological concerns around subjective, non-standardised outcome assessment, potentially non-representative populations of total Nipah infection survivors, and unknown generalisability to Nipah virus outbreaks in South Asia.

## Data Availability

All data produced in the present work are contained in the manuscript.

## Contributors

NS conceptualised the study. TZ and QW conducted the search and study selection. TZ and NS extracted data and conducted the risk of bias assessment. NS analysed the data and wrote the first draft of the manuscript. All authors were responsible for the methodology, data interpretation, review, and editing of the manuscript, and approved the final version of the manuscript. All authors had full access to all the data in the study and had final responsibility for the decision to submit for publication.

## Data sharing

Most extracted data from the included studies can be found in the manuscript and supplement; any additional data are available upon request to the corresponding author.

## Acknowledgments

NS acknowledges funding from the MRC Centre for Global Infectious Disease Analysis (reference MR/X020258/1), funded by the UK Medical Research Council (MRC). This UK funded award is carried out in the frame of the Global Health EDCTP3 Joint Undertaking. NS also acknowledges funding from Wellcome Trust (220900/Z/20/Z). The funders of the study had no role in study design, data collection, data analysis, data interpretation, or writing of the report.

