## Supplementary Materials for "Post-acute sequelae after Nipah virus infection: a systematic review"

### Supplementary methods

#### Risk of bias assessment

**Table S1. Modified JBI critical appraisal tool applied to each included study.** Details on the questions can be found in the checklist for prevalence, cohort and case-control studies.^1,2^

|  | Yes | No | Unclear | Not applicable |
| --- | --- | --- | --- | --- |
| **All studies** | | | | |
| 1. Was the sample frame appropriate to address the target population? | □ | □ | □ | □ |
| 2. Were study participants sampled in an appropriate way? | □ | □ | □ | □ |
| 3. Were the study subjects and the setting described in detail? | □ | □ | □ | □ |
| 4. Were valid methods used for the identification of the exposure (infection)? | □ | □ | □ | □ |
| 5. Were valid methods used for the identification of the outcome (post-acute condition)? | □ | □ | □ | □ |
| 6. Was the exposure (infection) measured in a standard, reliable way for all participants? | □ | □ | □ | □ |
| 7. Was the outcome (post-acute condition) measured in a standard, reliable way for all participants? | □ | □ | □ | □ |
| 8. Was the response rate adequate, and if not, was the low response rate managed appropriately? | □ | □ | □ | □ |
| 9. Were the participants free of the outcome at the start of the study (or before the infection)? | □ | □ | □ | □ |
| 10. Were outcomes to be extracted clearly described? | □ | □ | □ | □ |
| 11. Was the follow up time reported and sufficient to be long enough for outcomes to occur? | □ | □ | □ | □ |
| **Longitudinal studies** |  |  |  |  |
| 12. Was follow up complete, and if not, were the reasons to loss to follow up described and explored? | □ | □ | □ | □ |
| 13. Were strategies to address incomplete follow up utilized? | □ | □ | □ | □ |
| **Studies with control group** | | | | |
| 14. Were the two groups comparable and recruited from the same population? | □ | □ | □ | □ |
| 15. Were confounding factors identified and accounted for? | □ | □ | □ | □ |

#### Overlap in included articles

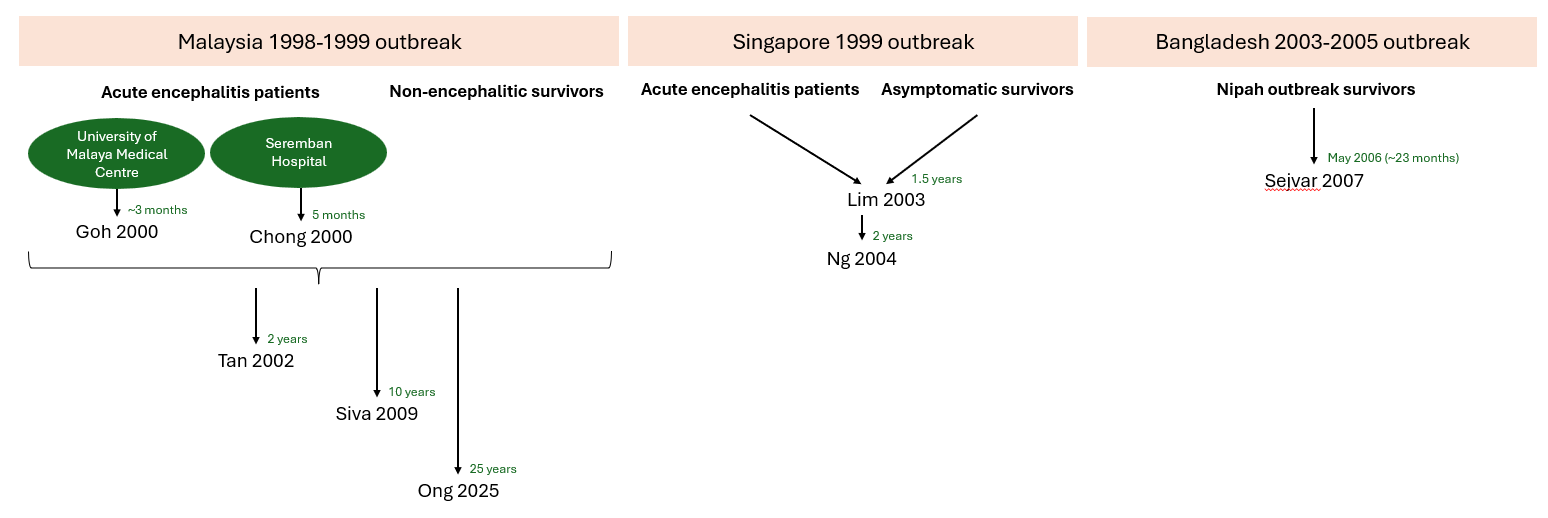
**Figure S1. Overview of included articles and study populations.** The end of follow-up/time at measurement after acute infection is indicated for each article.

In Malaysia, the Nipah virus outbreak occurred between September 1998 and June 1999. 265 patients who developed acute encephalitis were admitted to Malaysian hospitals nationwide. Goh 2000 followed all 94 Nipah patients hospitalised at the University of Malaya Medical Center,^3^ and Chong 2000 followed all 103 patients treated in Seremban Hospital (Figure S1).^4^ Tan 2002 followed all encephalitis survivors in the country (i.e. all participants in Goh 2000 and Chong 2000; other hospitals not shown on diagram) and additionally included 89 participants with non-encephalitic or asymptomatic infection.^5^ Siva 2009 likely covers a subset of participants in these previous studies, as they included 12 encephalitis and 27 non-encephalitic survivors identified via serology, only 7 of which had not previously been admitted to a hospital.^6^ Similarly, Ong 2025 recruited 25 patients with a clinical history of Nipah virus infection during the 1998 outbreak from Kampung Sungai Nipah, but their original location of hospitalisation was not specified.^7^

In Singapore, the first case of Nipah infection was detected in 1999. Lim 2003 followed all 11 hospitalised encephalitis patients in the country,^8^ and Ng 2004 reports a longer follow-up of the same patients (with a focus on different symptoms).^9^ Lim 2003 additionally included 9 abattoir workers with asymptomatic Nipah virus infection.

In Bangladesh, only one article was included (Sejvar 2007), which followed survivors of Nipah virus outbreaks in 2003, 2004 and 2005 regardless of their presentation during acute infection.^10^

#### Meta-analytic methods

Meta-analysis was conducted using the *meta* package version 8.0-2.^11^ For meta-analysis of proportions (prevalence of post-acute sequelae and cumulative risk of late-onset or relapsing neurological symptoms after the initial Nipah infection), a logit transformation was applied and estimates were pooled using a random intercept logistic regression model.^12,13^ The maximum likelihood estimator was used to calculate the absolute between-study variance *τ^2^*. Publication bias for the prevalence of neurological deficit was assessed qualitatively using a funnel plot, but the number of included studies was too small for a quantitative evaluation.

To further investigate encephalitis during acute infection as a risk factor for post-acute sequelae among Nipah survivors, we conducted a random-effects meta-analysis of prevalence ratios using the generic inverse variance method and the restricted maximum likelihood between-study variance estimator. A continuity correction of 0.5 was used in cases of zero counts to estimate the log prevalence ratio.

All meta-analyses included few studies per subgroup, which is associated with imprecise estimates of between-study variance.^14^ We therefore compared 95% confidence intervals around the pooled estimate derived using the Hartung-Knapp adjustment and the DerSimonian and Laird method. We used the Hartung-Knapp method for calculation of confidence intervals unless this led to non-informative estimates.^14^ Between-study heterogeneity in the meta-analyses was assessed qualitatively through visual inspection of the forest plots. We also calculated *τ^2^*, *I^2^* and 95% prediction intervals. Prediction intervals were based on the t-distribution with *k-1* degrees of freedom and reduce to the confidence interval if *τ^2^* is 0.^15^ Due to the small number of studies, it was not possible to further investigate potential sources of heterogeneity.

### Supplementary results

#### Description of excluded studies

Two additional studies and two case reports did not meet eligibility criteria, but provide some information on potential residual neurological deficits. In a report of the 2023 Nipah outbreak in Kerala, India, none of 4 survivors of encephalitis experienced residual neurological deficits (or respiratory symptoms) at time of hospital discharge but it is not known whether any patients subsequently developed late-onset sequelae.^16^ In a 2014 henipavirus outbreak in the Philippines, 2 of 8 (25%) survivors of infection overall and 2 of 2 (100%) survivors of encephalitis had residual neurological symptoms (severe cognitive impairment, motor weakness, ataxia and ophthalmoplegia) after a mean follow-up of 50 days (range 31-75 days) after acute symptom onset.^17^ In an excluded case report otherwise meeting inclusion criteria, a Nipah encephalitis survivor from a 2020 Bangladesh outbreak experienced persistent neurological and psychiatric sequelae 17 months after acute infection, though her symptoms reduced in severity and intensity over time.^18^ Another case report describes a potential case of late-onset encephalitis with a long delay.^19^ A woman with exposure to Nipah virus during the 1998-1999 outbreak in Malaysia was asymptomatic, but developed Nipah virus antibodies and characteristic brain lesions. She experienced encephalitis 11 years later, with a relapse the following year, which could not be attributed to other causes.

#### Risk of bias assessment

**Table S2. Risk of bias assessment.** A percentage of “Yes” responses among applicable items of 0-49%, 50-74% and 75-100% were defined as representing low, moderate and high quality, respectively. Recruitment of all hospitalised patients in studies on Nipah encephalitis was considered appropriate sampling. In Siva, Sejvar and Ong, a convenience sample of non-encephalitic survivors seems to have been taken, though recruitment was not otherwise described. In the earliest two articles, the case definition for Nipah virus infection was based on epidemiological and clinical features, which was considered valid in the absence of readily available confirmatory tests. All but one of the later articles included serological confirmation. Post-acute sequelae were usually assessed through physical and/or neurological examination. Three studies (Siva, Sejvar, Ong) additionally developed a subjective symptom questionnaire, and one study (Ng) included a psychiatric evaluation and assessed cognitive impairment using neuropsychological testing. U = Unclear, NA = not applicable.

|  | **Goh 2000** | **Chong 2000** | **Tan 2002** | **Siva 2009** | **Ong 2025** | **Lim 2003** | **Ng 2004** | **Sejvar 2007** |
| --- | --- | --- | --- | --- | --- | --- | --- | --- |
| 1. Appropriate sample frame | Yes | Yes | Yes | Yes | Yes | Yes | Yes | Yes |
| 2. Appropriate sampling | Yes | Yes | Yes | No | No | U | Yes | No |
| 3. Detailed description of participants and setting | Yes | Yes | No | Yes | Yes | Yes | Yes | Yes |
| 4. Valid methods for identification of infection | Yes | Yes | Yes | Yes | No | Yes | Yes | Yes |
| 5. Valid methods for identification of post-acute outcome | U | U | U | Yes | U | U | Yes | Yes |
| 6. Standard and reliable measurement of infection | Yes | Yes | Yes | Yes | U | Yes | Yes | Yes |
| 7. Standard and reliable measurement of post-acute outcome | U | U | U | No | Yes | U | Yes | Yes |
| 8. Adequate response rate | Yes | Yes | U | U | U | U | Yes | Yes |
| 9. Participants confirmed to be free of post-acute conditions before infection | Yes | U | No | No | No | Yes | No | Yes |
| 10. Clear description of outcomes | No | Yes | Yes | Yes | Yes | Yes | Yes | Yes |
| 11. Sufficient follow-up duration | U | Yes | Yes | Yes | Yes | Yes | Yes | Yes |
| 12. Complete follow-up or reasons for loss described and explored | U | Yes | Yes | NA | NA | No | No | Yes |
| 13. Strategies to address incomplete follow-up | U | NA | NA | NA | NA | No | No | NA |
| 14. Groups recruited from the same population | NA | NA | NA | Yes | NA | NA | NA | NA |
| 15. Confounding factors identified and accounted for | NA | NA | NA | No | NA | NA | NA | NA |
| Percentage “Yes” | 54% | 75% | 58% | 62% | 45% | 54% | 77% | 92% |
| Overall appraisal | Include | Include | Include | Include | Include | Include | Include | Include |

#### Meta-analyses

**Table S3. Extraction decisions for the prevalence of residual neurological deficits in Nipah survivors.**

| **Study** | **Numerator** | **Denominator** | **Decision** | **Residual neurological deficits** | **Sensitivity analysis** |
| --- | --- | --- | --- | --- | --- |
| **Total survivors of Nipah infection** | | | | | |
| Lim 2003 | 4 | 20 | The denominator includes 11 encephalitis survivors with known sequelae status (1 of 13 died and 1 was lost to follow-up before assessment at 18 months), and 9 asymptomatic infection survivors (all with known sequelae status).  The numerator includes 4 cases of residual neurological deficit among encephalitis survivors (Table 1) and 0 cases among asymptomatic infection survivors (“All nine asymptomatic subjects (all men, mean age 50.1 years) were clinically well during the outbreak and follow up.”). | Focal weakness, dysaesthesia, Horner syndrome, monoparesis, nystagmus, cranial nerve palsy, branch retinal artery occlusion (cognitive impairment not assessed) | None |
| Siva 2009 | 8 | 39 | The denominator includes the 39 identified seropositive Nipah virus infection survivors, which all underwent detailed follow-up neurological examinations. 12 of 39 had Nipah encephalitis and 27 of 39 had had non-encephalitic acute infection, as shown in Table 1.  The numerator includes 8 cases of “significant” persistent neurological deficit among Nipah encephalitis survivors, shown in Table 2, and 0 cases among non-encephalitic survivors. The article states: “All asymptomatic Nipah infection survivors had no neurological deficit. Significant neurological sequelae were only found in survivors of Nipah encephalitic.” | Vegetative state, tetraparesis, cranial nerve palsy, tinnitus, limb dysmetria and dysdiadochokinesia, nystagmus, dysarthria, cognitive impairment | None |
| Sejvar 2007 | 7 | 21 | This article reported outcomes at two timepoints [FU1 in July-August 2005 (median 14 months after acute infection) and FU2 in May 2006]. We extracted the datapoint at the later follow-up, but prevalence was very similar at both (7/22 = 32% at FU1 vs. 7/21 = 33% at FU2).  The denominator includes all 21 Nipah infection survivors with sequelae assessment at the second follow-up (1 patient was lost to follow-up between FU1 and FU2). This is made up of 17 patients with encephalitis during acute infection and 4 patients with non-encephalitic “acute febrile illness”. The patient lost to follow-up had acute febrile illness.  The numerator includes the 7 encephalitis survivors with “moderate-to-severe” neurological dysfunction (“Seven patients (32%), all recovering from encephalitis, had moderate-to-severe objective neurological dysfunction that had persisted since, or developed subsequent to, onset of acute NiV encephalitis.”). These 7 cases are later referred to as “persistent neurological abnormalities”, and were observed at both follow-up assessments.  We extracted data for cases referred to as “moderate-to-severe objective neurological dysfunction”/”persistent neurological abnormalities”, as this was most similar to descriptions of residual neurological deficits in other papers. Additionally, alternative statements in the paper were unclear, and the necessary details in the supplement could not be retrieved. Specifically, the authors state that: “In 12 patients, neurological examination was normal at both evaluations (see Supplementary Table); the patient not assessed at FU2 also had a normal neurological examination.” This implies that neurological examination was non-normal in 10 of 22 patients at FU1 and in 9 of 21 patients at FU2. It is nevertheless unclear whether these numbers include, or are additional to, 2 pre-existing or age-related cases (“One patient, aged 50, had pre-existing mild bradykinesia; a second patient, aged 7, had mild gait imbalance reflective of a normal variation for age.”)  Alternative methodological decisions could have resulted in the following numbers: 7/22 (32%), 10/22 (45%), 9/21 (43%). | Cognitive dysfunction/developmental delay, ataxia/gait abnormalities, focal weakness, cervical dystonia, dysarthria, nystagmus, bradykinesia, difficulty with rapid movements | None |
| **Survivors of Nipah encephalitis** | | | | | |
| Chong 2000 | 20 | 61 | The article followed 103 encephalitis patients treated in hospital, of which 41% were reported to have died. The denominator therefore includes the remaining 61 survivors.  There are discrepancies in reporting of the numerator in the article. The results state that: “When the surviving patients were last seen 5 months since the outbreak, some 40% recovered fully while 19% had residual neurological deficits which were: cerebellar signs (6.8%), tetraparesis (4.9%), cranial nerve palsies (3.9%), monoparesis (1.0%), peripheral nerve lesions (2.9%) and higher mental function deficits (2.9%). Of the 4 patients with cranial nerve palsies, […]”. As 4 patients with cranial nerve palsies represent 3.9% of the total, this implies that these numbers apply to the total number of encephalitis patients rather than just the survivors, as the writing would suggest. Therefore, we extracted the numerator with residual neurological deficits as 19% of 103, i.e. 20. However, this is not consistent with the following statement in the discussion: “Most of the 23 patients (19%) with residual neurological deficits were mild.” | Cerebellar signs, tetraparesis, cranial nerve palsy, monoparesis, peripheral nerve lesions, medial temporal syndrome, dysphasia, vegetative state | Exchanged for Siva 2009 in Figure S3A. Chong 2000 was included over Siva 2009 due to a larger sample size. |
| Ng 2004 | 7 | 9 | The denominator includes the 9 Nipah encephalitis survivors followed in the article (of 13 patients affected by the outbreak in Singapore, 1 died and 2 were lost to follow-up).  The numerator includes the 7 survivors with residual neurological deficit and/or cognitive impairment shown in Table 1. | Focal weakness, dysaesthesia, Horner syndrome, monoparesis, nystagmus, cranial nerve palsy, branch retinal artery occlusion, cognitive impairment | Exchanged for Lim 2003 in Figure S3C. Ng 2004 was included over Lim 2003 due to the inclusion of cognitive impairments among neurological deficits (consistent with the other studies). The prevalence in Lim 2003 is lower as a result. |
| Sejvar 2007 | 7 | 17 | See above | See above | None |
| Goh 2000 | 14 | 64 | This article was excluded from the primary meta-analysis as alignment with our case definition for post-acute sequelae (minimum 3 months persistence) could not be confirmed.  The denominator includes 64 Nipah encephalitis survivors.  The numerator includes 14 patients with residual neurological deficits (“Thirty patients (32 percent) died, 50 patients (53 percent) recovered fully, and 14 patients (15 percent) had residual neurologic deficits.”). | Vegetative state, cognitive impairment, cognitive and cerebellar disabilities | Included in Figure S4 |
| **Survivors of late-onset or relapsed Nipah encephalitis** | | | | | |
| Tan 2002 | 11 | 18 | The article follows 12 patients with relapsed encephalitis and  10 patients with late-onset encephalitis, 4 of which died. The denominator includes the 18 survivors in this cohort shown in Table 1.  The numerator includes the 11 cases of residual neurological deficits shown in Table 1 (column “Outcome”). | Ataxia, cognitive impairment, cranial nerve palsy, nystagmus, dysphasia, tetraparesis, epilepsy | None |

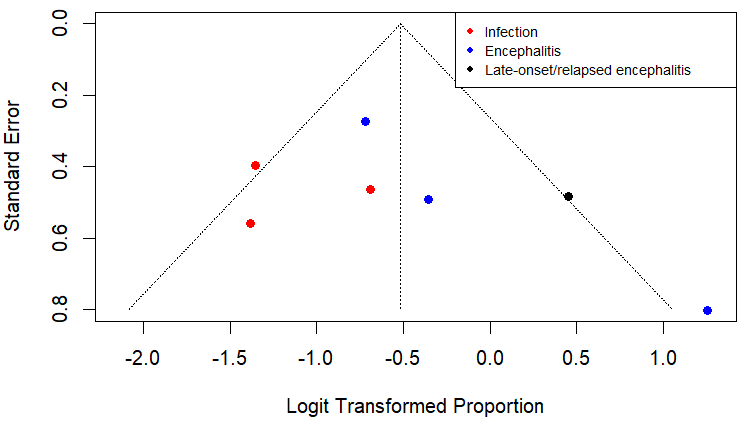

**Figure S2. Funnel plot for pooled prevalence of residual neurological deficits in Nipah survivors.** Individual estimates are shown for the different subgroups (total Nipah infection survivors, Nipah encephalitis survivors and late-onset/relapsed Nipah encephalitis survivors).

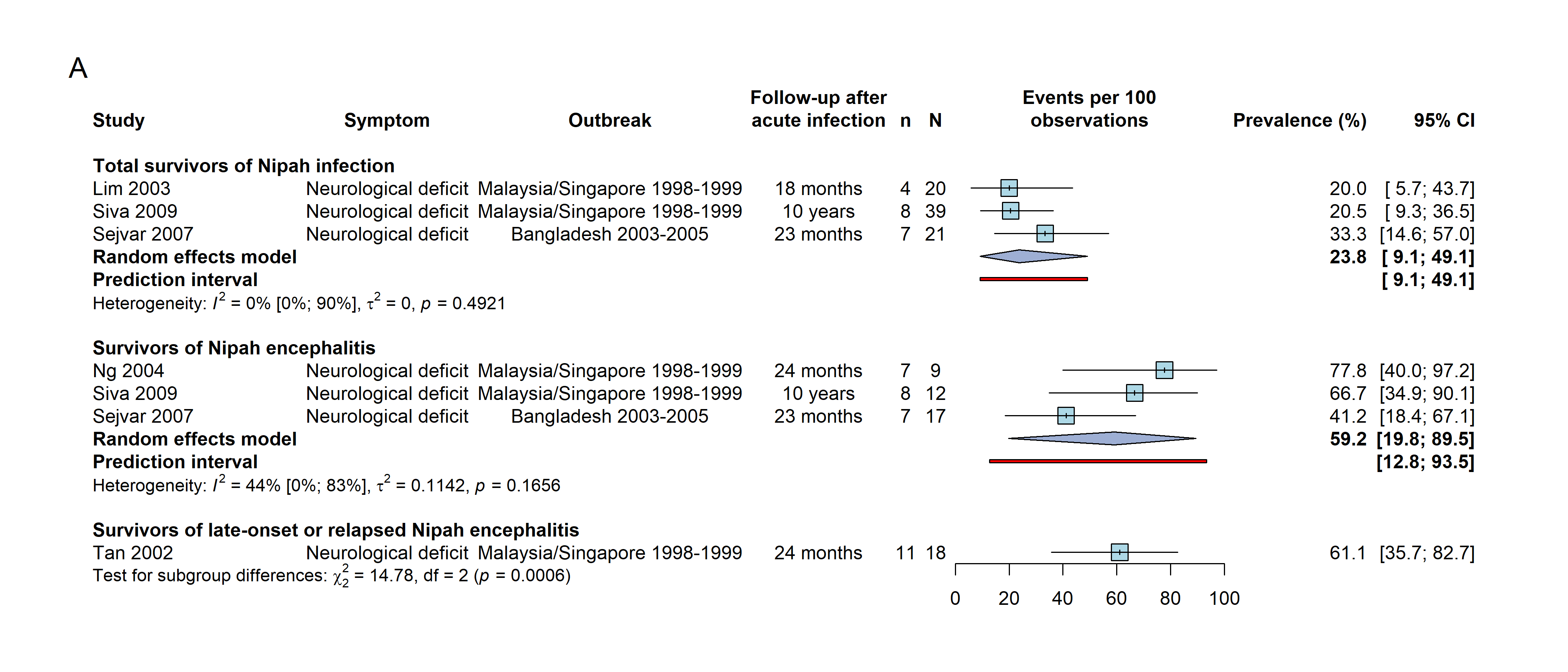

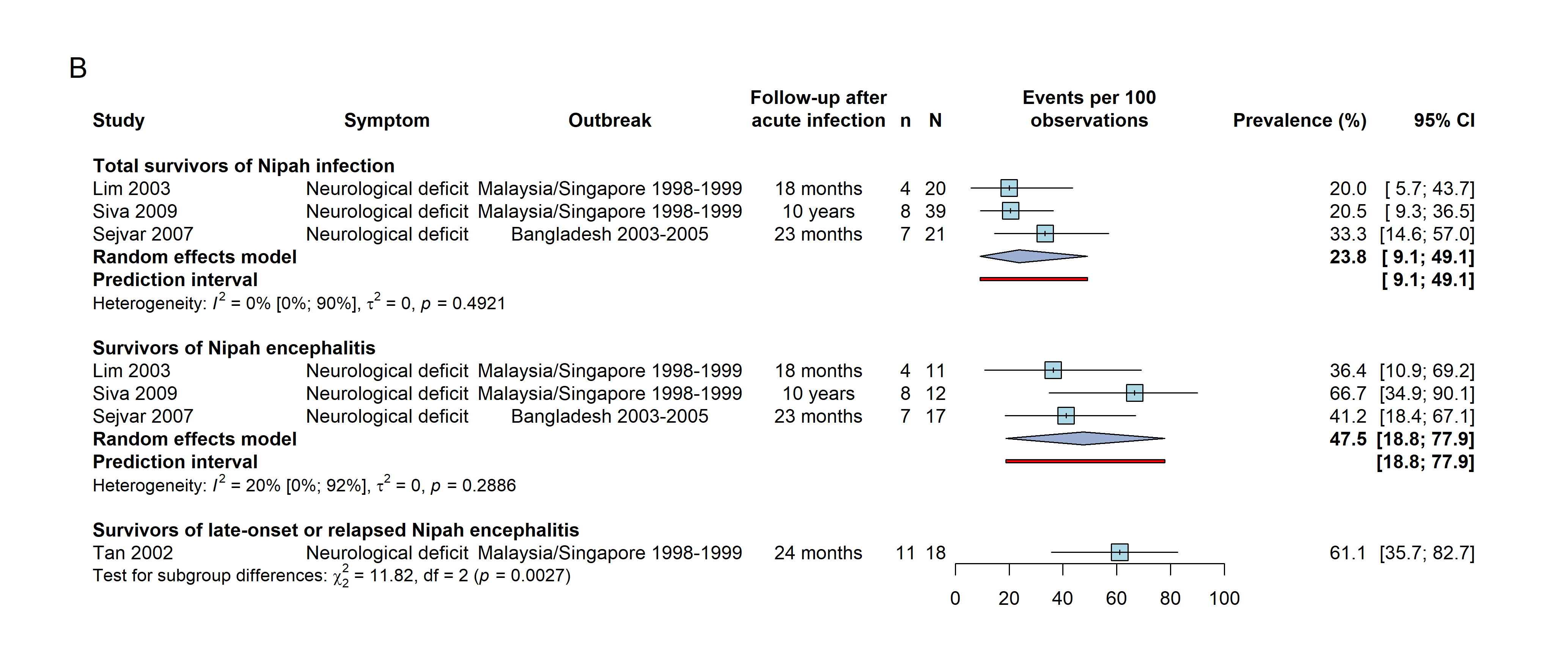

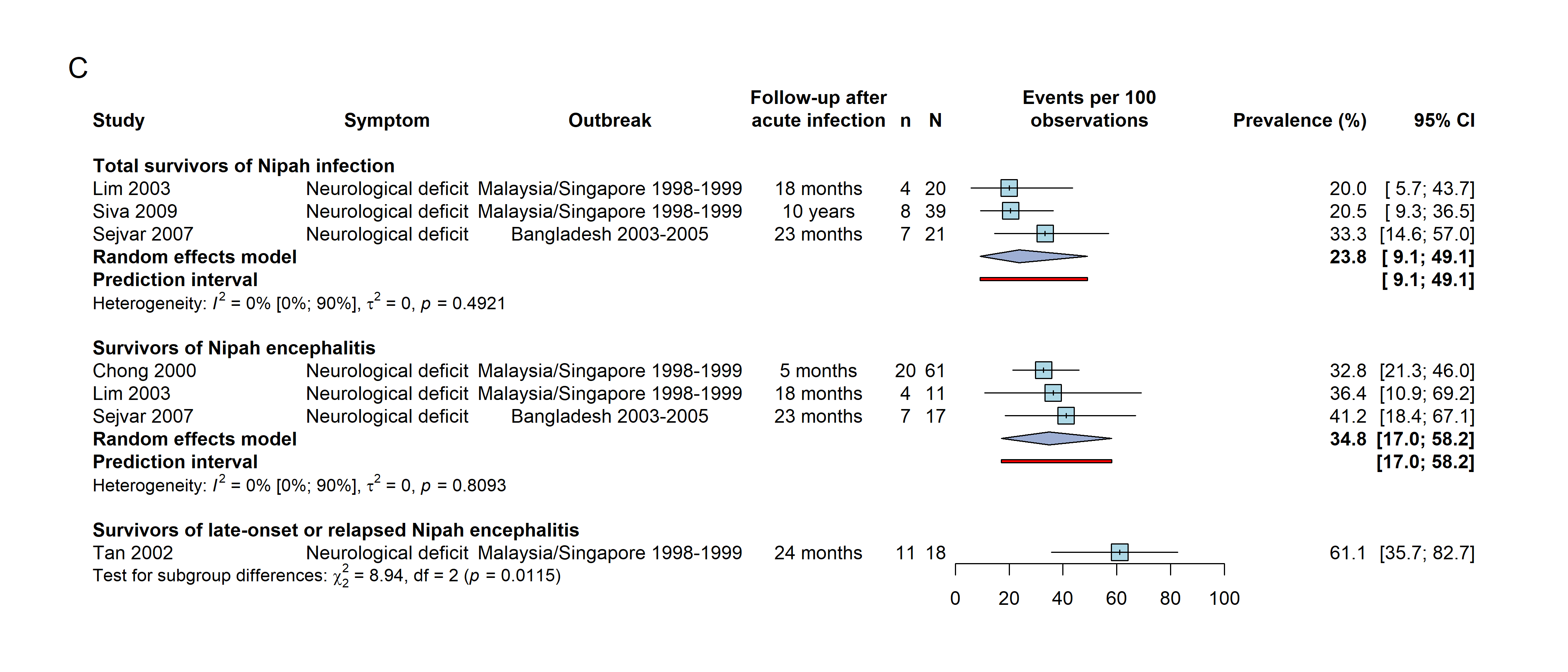

**Figure S3. Sensitivity analysis for random-effects meta-analysis of the prevalence of residual neurological deficits after infection with Nipah virus.** In each panel, prevalence estimates are shown for three subgroups: total survivors of Nipah infection, survivors of Nipah encephalitis (a subset of all survivors of Nipah infection), and survivors of late-onset or relapsed Nipah encephalitis (one study). In panels A-C, different articles were included for the estimate among survivors of Nipah encephalitis. Ng 2004 and Lim 2003, and Siva 2009 and Chong 2000, partially included the same patients and were therefore not both included in the meta-analysis at the same time.

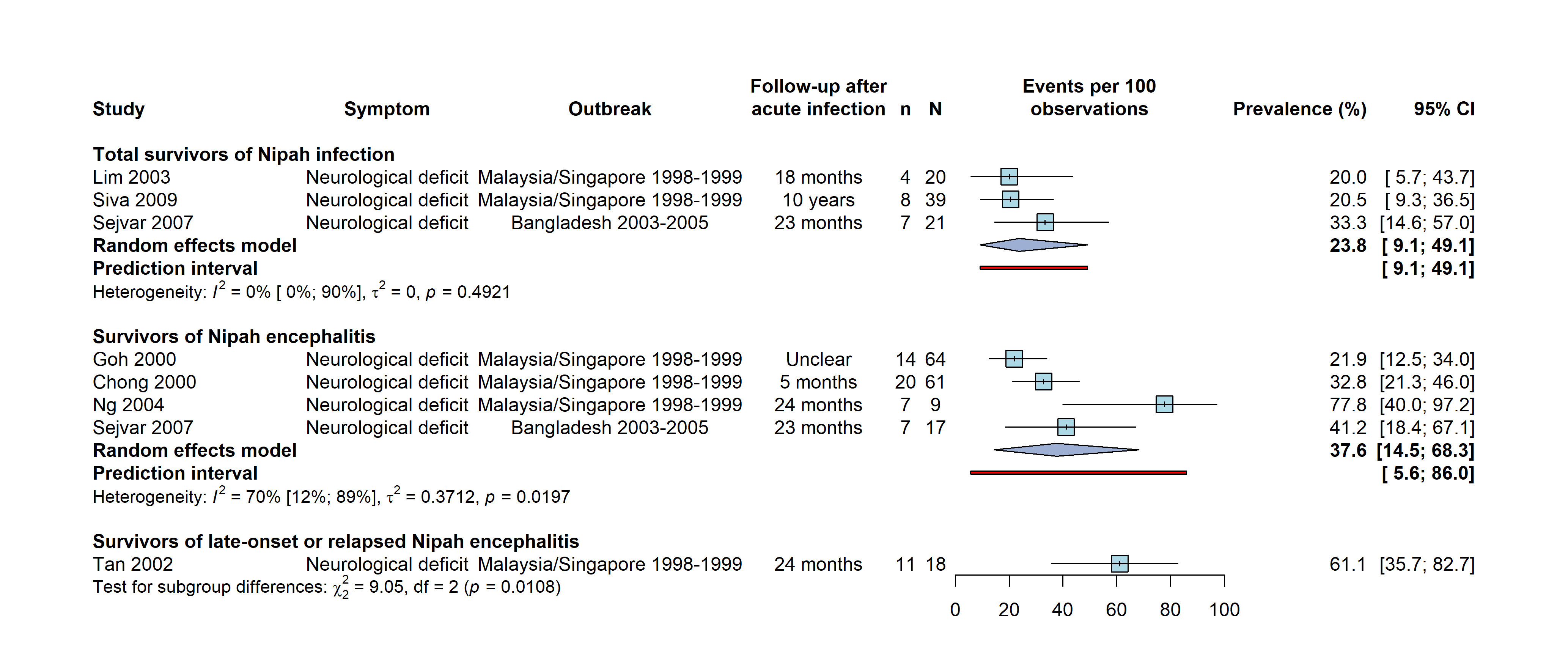

**Figure S4. Sensitivity analysis for random-effects meta-analysis of the prevalence of residual neurological deficits after infection with Nipah virus.** Prevalence estimates are shown for three subgroups: total survivors of Nipah infection, survivors of Nipah encephalitis (a subset of all survivors of Nipah infection), and survivors of late-onset or relapsed Nipah encephalitis (one study). Compared to the primary analysis, the estimate among survivors of Nipah encephalitis here includes Goh 2000, which was excluded because the follow-up time was not clearly specified and so the case definition for post-acute sequelae might not be met.

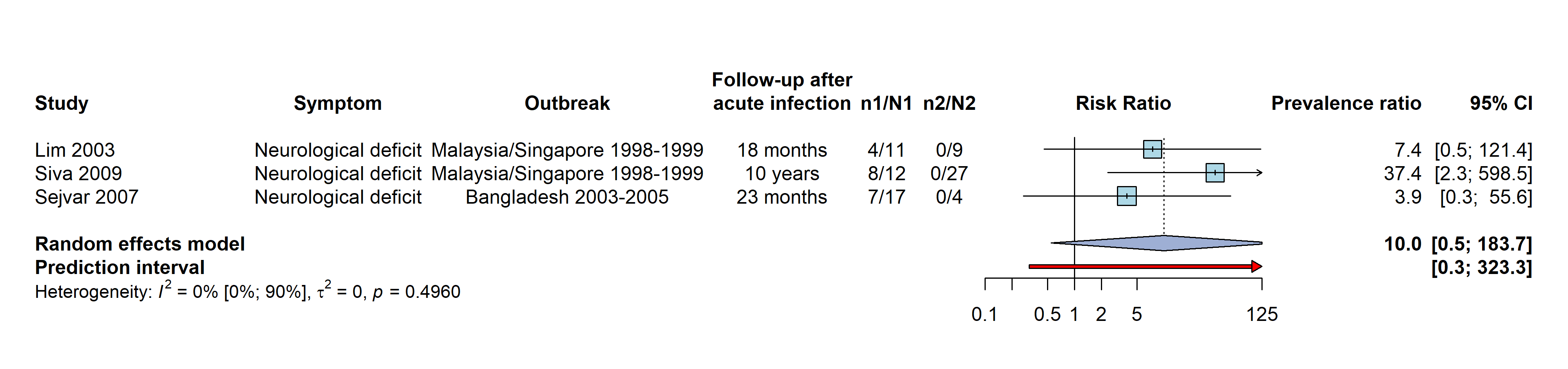
**Figure S5. Forest plot for prevalence ratio of neurological deficit in survivors of Nipah encephalitis compared to survivors with non-encephalitic acute infection.** 95% confidence intervals for the pooled estimate were calculated using the Hartung-Knapp method.

**Table S4: Random-effects meta-analysis for the prevalence of individual post-acute symptoms.** For each symptom, the pooled prevalence was calculated for total Nipah infection survivors and for Nipah encephalitis survivors. 95% confidence intervals around pooled estimates were calculated using the DerSimonian and Laird method.

|  | **Prevalence (%)  (95% confidence interval)** | ***τ^2^*** | ***I^2^* (%)** | **95% prediction interval (%)** | **p-value for subgroup difference** | **Studies** |
| --- | --- | --- | --- | --- | --- | --- |
| **Cognitive impairment** | | | | | | |
| • Nipah infection | 11.7 (5.7-22.5) | 0.0 | 38 | 0.08-95.6 | 0.05 | Siva 2009, Sejvar 2007 |
| • Nipah encephalitis | 45.9 (14.1-81.5) | 0.9 | 81 | 0.0-100.0 |  | Ng 2004, Sejvar 2007 |
| **Ataxia** | | | | | | |
| • Nipah infection | 5.0 (1.9-12.6) | 0.0 | 0 | 0.6-32.4 | 0.30 | Lim 2003, Siva 2009, Sejvar 2007 |
| • Nipah encephalitis | 10.7 (3.5-28.4) | 0.0 | 0 | 0.0-99.7 |  | Lim 2003, Sejvar 2007 |
| **Cranial nerve palsy** | | | | | | |
| • Nipah infection | 7.0 (2.2-19.8) | 0.4 | 54 | 0.2-77.9 | 0.97 | Lim 2003, Siva 2009, Sejvar 2007 |
| • Nipah encephalitis | 7.1 (3.0-16.0) | 0.0 | 0 | 0.0-96.6 |  | Chong 2000, Ng 2004 |
| **Nystagmus** | | | | | | |
| • Nipah infection | 7.0 (2.2-19.8) | 0.4 | 54 | 0.2-77.9 | 0.14 | Lim 2003, Siva 2009, Sejvar 2007 |
| • Nipah encephalitis | 19.2 (8.2-38.7) | 0.0 | 0 | 0.0-99.2 |  | Ng 2004, Sejvar 2007 |
| **Monoparesis** | | | | | | |
| • Nipah encephalitis | 2.9 (0.7-10.7) | 0.0 | 47 | 0.0-99.6 | / | Chong 2000, Ng 2004 |
| **Dysarthria** | | | | | | |
| • Nipah infection | 3.3 (0.8-12.4) | 0.0 | 0 | 0.0-99.7 | 0.46 | Siva 2009, Sejvar 2007 |
| • Nipah encephalitis | 6.9 (1.7-23.8) | 0.0 | 0 | 0.0-99.9 |  | Siva 2009, Sejvar 2007 |
| **Focal weakness** | | | | | | |
| • Nipah infection | 7.3 (2.4-20.4) | 0.0 | 0 | 0.0-99.4 | 0.56 | Lim 2003, Sejvar 2007 |
| • Nipah encephalitis | 11.5 (3.8-30.3) | 0.0 | 0 | 0.0-99.7 |  | Ng 2004, Sejvar 2007 |
| **Fatigue** | | | | | | |
| • Nipah infection (primary analysis) | 48.1 (23.4-73.8) | 0.5 | 87 | 0.0-100.0 | 0.57 | Siva 2009, Sejvar 2007 (at 6 months) |
| • Nipah infection (sensitivity analysis) | 25.0 (15.7-37.4) | 0.0 | 47 | 0.8-93.6 | 0.52 | Siva 2009, Sejvar 2007 (at 2 years) |
| • Nipah encephalitis | 35.4 (11.9-69.1) | 1.0 | 78 | 0.3-99.2 |  | Ng 2004, Siva 2009, Sejvar 2007 |

#### Prevalence data for all individual sequelae

**Table S5: Prevalence for all individual post-acute symptoms reported among total Nipah infection survivors and Nipah encephalitis survivors.** For symptoms reported in at least two independent studies, prevalence represents the weighted percentage from random-effects meta-analysis (*). For estimates from different non-independent articles, both values are shown.

|  | **Total survivors of Nipah infection** | | | | **Survivors of Nipah encephalitis** | | | |
| --- | --- | --- | --- | --- | --- | --- | --- | --- |
| **Symptom** | **Studies** | **n** | **N** | **Prevalence (%) (95% CI)** | **Studies** | **n** | **N** | **Prevalence (%) (95% CI)** |
| **Neurological symptoms: cognitive dysfunction** | | | | | | | | |
| Memory impairment | Sejvar | 5 | 21 | 23.8 (8.2-47.2) | / | / | / | / |
| Cognitive impairment | Siva, Sejvar | 7 | 60 | 11.7 (5.7-22.5)* | Ng, Sejvar | 10 | 25 | 45.9 (14.1-81.5)* |
| **Neurological symptoms: motor or sensory dysfunction** | | | | | | | | |
| Bradykinesia | Sejvar | 3 | 21 | 14.3 (3-36.3) | Sejvar | 3 | 17 | 17.6 (3.8-43.4) |
| Myoclonus | Sejvar | 3 | 21 | 14.3 (3-36.3) | / | / | / | / |
| Focal weakness | Lim, Sejvar | 3 | 41 | 7.3 (2.4-20.4)* | Ng, Sejvar | 3 | 26 | 11.5 (3.8-30.3)* |
| Cranial nerve palsy | Lim, Siva, Sejvar | 6 | 80 | 7.0 (2.2-19.8)* | Chong, Ng | 5 | 70 | 7.1 (3.0-16.0)* |
| Nystagmus | Lim, Siva, Sejvar | 6 | 80 | 7.0 (2.2-19.8)* | Ng, Sejvar | 5 | 26 | 19.2 (8.2-38.7)* |
| Ataxia | Lim, Siva, Sejvar | 4 | 80 | 5.0 (1.9-12.6)* | Lim, Sejvar | 3 | 28 | 10.7 (3.5-28.4)* |
| Branch retinal artery occlusion | Lim | 1 | 20 | 5.0 (0.1-24.9) | Lim/Ng | 1/1 | 11/9 | 9.1 (0.2-41.3)/ 11.1 (0.3-48.2) |
| Dysaesthesia | Lim | 1 | 20 | 5.0 (0.1-24.9) | Lim/Ng | 1/1 | 11/9 | 9.1 (0.2-41.3)/ 11.1 (0.3-48.2) |
| Horner syndrome | Lim | 1 | 20 | 5.0 (0.1-24.9) | Lim/Ng | 1/1 | 11/9 | 9.1 (0.2-41.3)/ 11.1 (0.3-48.2) |
| Monoparesis | Lim | 1 | 20 | 5.0 (0.1-24.9) | Chong, Ng | 2 | 70 | 2.9 (0.7-10.7)* |
| Cervical dystonia | Sejvar | 1 | 21 | 4.8 (0.1-23.8) | Sejvar | 1 | 17 | 5.9 (0.1-28.7) |
| Dysarthria | Siva, Sejvar | 2 | 60 | 3.3 (0.8-12.4)* | Siva, Sejvar | 2 | 29 | 6.9 (1.7-23.8)* |
| Abnormal movement | Siva | 1 | 39 | 2.6 (0.1-13.5) | Siva | 1 | 12 | 8.3 (0.2-38.5) |
| Dysdiadochokinesia | Siva | 1 | 39 | 2.6 (0.1-13.5) | / | / | / | / |
| Numbness | Siva | 1 | 39 | 2.6 (0.1-13.5) | Siva | 1 | 12 | 8.3 (0.2-38.5) |
| Tetraparesis | Siva | 1 | 39 | 2.6 (0.1-13.5) | Chong | 5 | 61 | 8.2 (2.7-18.1) |
| Cerebellar dysfunction | / | / | / | / | Chong | 7 | 61 | 11.5 (4.7-22.2) |
| Peripheral nerve lesions | / | / | / | / | Chong | 3 | 61 | 4.9 (1-13.7) |
| **Neurological symptoms: other** | | | | | | | | |
| Seizures/epilepsy | Siva | 3 | 39 | 7.7 (1.6-20.9) | Siva | 2 | 12 | 16.7 (2.1-48.4) |
| Reduced consciousness/vegetative state | Siva | 1 | 39 | 2.6 (0.1-13.5) | Chong/Siva | 1/1 | 61/12 | 1.6 (0-8.8)/ 8.3 (0.2-38.5) |
| Dysphasia | / | / | / | / | Chong | 1 | 61 | 1.6 (0-8.8) |
| Medial temporal syndrome | / | / | / | / | Chong | 1 | 61 | 1.6 (0-8.8) |
| **Psychiatric symptoms** | | | | | | | | |
| Behaviour/personality changes | Sejvar | 4 | 21 | 19.0 (5.4-41.9) | Ng | 2 | 9 | 22.2 (2.8-60) |
| Irritability | Siva | 2 | 39 | 5.1 (0.6-17.3) | Siva | 1 | 12 | 8.3 (0.2-38.5) |
| Depression | Lim | 1 | 20 | 5.0 (0.1-24.9) | Lim/Ng | 1/5 | 11/9 | 9.1 (0.2-41.3)/ 55.6 (21.2-86.3) |
| **Systemic & other symptoms** | | | | | | | | |
| Fatigue | Siva, Sejvar | 27 | 61 | 48.1 (23.4-73.8)* [primary analysis] | Ng, Siva, Sejvar | 16 | 38 | 35.5 (11.9-69.1)* |
| Daytime somnolence | Siva | 10 | 39 | 25.6 (13-42.1) | Siva | 3 | 12 | 25.0 (5.5-57.2) |
| Headache | Siva | 6 | 39 | 15.4 (5.9-30.5) | Siva | 1 | 12 | 8.3 (0.2-38.5) |
| Dizziness | Siva | 3 | 39 | 7.7 (1.6-20.9) | Siva | 0 | 12 | 0.0 (0-26.5) |
| Cough | Siva | 1 | 39 | 2.6 (0.1-13.5) | Siva | 0 | 12 | 0.0 (0-26.5) |
| Tinnitus | Siva | 1 | 39 | 2.6 (0.1-13.5) | / | / | / | / |
| Weight gain | Siva | 1 | 39 | 2.6 (0.1-13.5) | Siva | 0 | 12 | 0.0 (0-26.5) |

#### Association of post-acute sequelae with Nipah

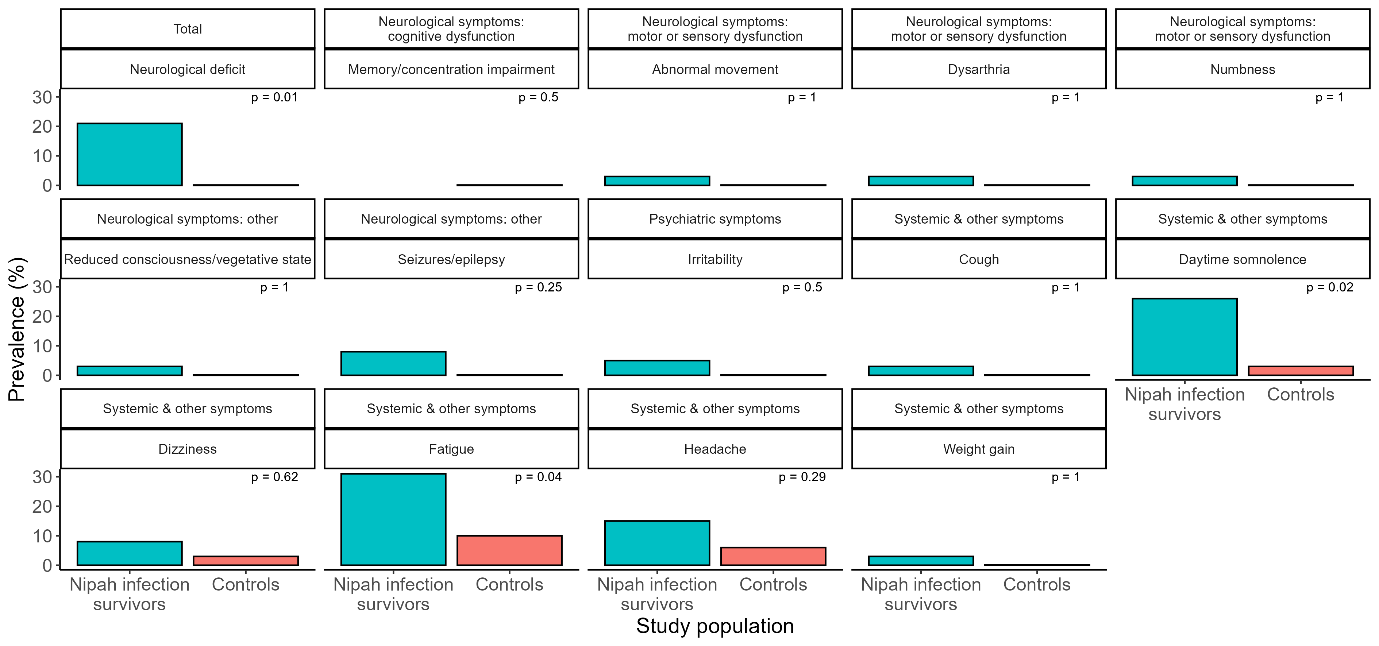

**Figure S6. Bar chart of prevalence of different post-acute symptoms among total Nipah infection survivors and a control group without past Nipah infection in Siva et al.^6^** The p-value from Fisher’s exact test for the association of Nipah infection status with the post-acute symptom is shown in each panel.

#### Late-onset neurological sequelae

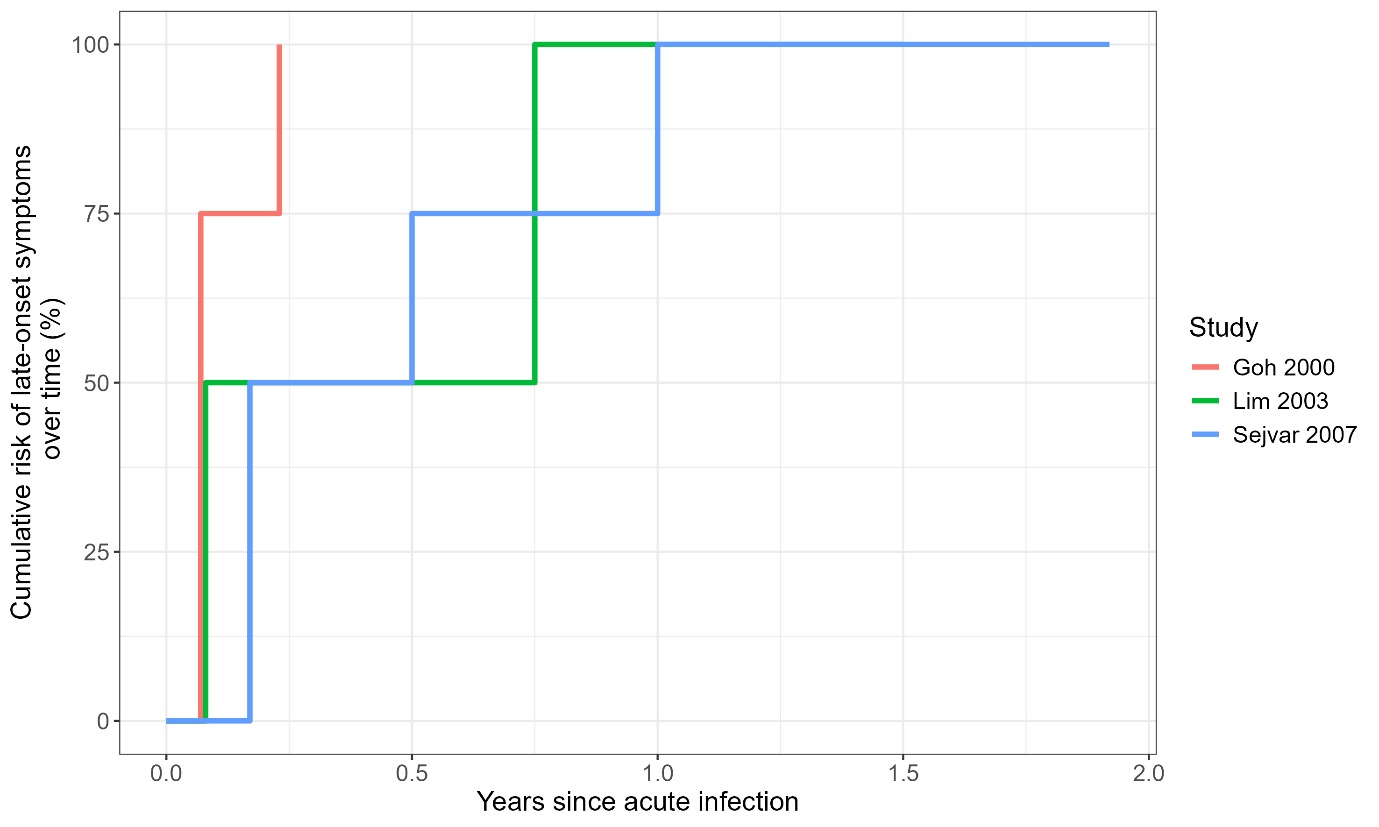

**Figure S7. Timing of late-onset neurological sequelae among all affected patients.** Note that total follow-up time in Goh 2000 was not known.

#### Functional outcomes and disability

**Table S6. Descriptions of functional outcomes associated with residual neurological deficits in the included studies.**

| **Study** | **Description** |
| --- | --- |
| Goh 2000 | Of 14 patients with residual neurological deficits (unclear timing), 5 (36%) were in a persistent vegetative state, 2 (14%) had residual cognitive impairment and were dependent on caregivers, and 5 (36%) had mild disability (others unknown). |
| Chong 2002 | Of 20 patients with neurological deficits 5 months after the infection, 1 (5%) was in a persistent vegetative state, 2 (10%) had marked disabilities and were dependent on caregivers, and 4 (20%) had mild disabilities and were able to live independently (others unknown). |
| Ng 2004 | Of 9 encephalitis survivors at 24 months after infection, 7 (78%) were employed 2 years later (4 of which resumed their previous job) and 2 (22%) were unemployed due to disability. |
| Siva 2009 | Functional outcomes were assessed using the Modified Rankin Scale.  Of 13 Nipah encephalitis survivors at 10 years after infection, 3 (23%) had no symptoms/disability (score 0), 4 (31%) had no significant disability (score 1, able to carry out all usual activities despite some symptoms), 7 (54%) had slight disability (score 2, able to look after own affairs without assistance, but unable to carry out all previous activities) and 2 (15%) had moderate to severe disability (score 3-5, requiring help, from being able to walk unassisted to requiring constant nursing care).  Of 27 asymptomatic or non-encephalitic survivors at 10 years after infection, 13 (48%) had no symptoms/disability (score 0), 14 (52%) had no significant disability (score 1), 0 had slight disability (score 2) and 0 had moderate to severe disability (score 3-5). |
| Sejvar 2007 | Functional outcomes were assessed using a Functional Self-Assessment Rating Scale covering different domains (overall functioning, performance at work/school, housework/daily chores, mobility (walking, climbing stairs/ladder, and so forth), sleeping, thinking/concentration, memory, and mood/behaviour).  Among 17 Nipah encephalitis survivors and 5 febrile non-encephalitic survivors, 30% and 100% returned to normal functioning (at or above the level before the acute infection) by 23 months, respectively.  The median duration of time off work or school was 3 months (range 1 week to 2 years), and 2 of 18 patients were unemployed at 23 months. |
